# Beyond the Kidney: Extra-Renal Manifestations of Monogenic Nephrolithiasis and Their Significance

**DOI:** 10.1101/2023.05.16.23289588

**Authors:** Jad Badreddine, Kimberly Tay, Hsin-Ti Cindy Lin, Stephen Rhodes, Fredrick R. Schumacher, Donald Bodner, Chen-Han Wilfred Wu

## Abstract

**Objective:** To explore the frequency of occurrence of extra-renal manifestations associated with monogenic kidney stone diseases.

**Methods:** A literature review was conducted to identify genes that are well-established monogenic causes of nephrolithiasis. The Online Mendelian Inheritance in Man (OMIM) and Human Protein Atlas (HPA) databases were used to identify associated diseases and their properties. Disease phenotypes were ascertained using OMIM clinical synopses and sorted into 24 different phenotype categories as classified in OMIM. Disease phenotypes caused by the same gene were merged into a single gene-associated phenotype (GAP) unit such that one GAP encompasses all related disease phenotypes for a specific gene. We measured the total number of GAPs involving each phenotype category and determined the median phenotype category. Phenotype categories were classified as overrepresented or underrepresented if the number of GAPs involving them was higher or lower than the median, respectively. A chi-square test was conducted to determine whether the number of GAPs affecting a given category significantly deviated from the median.

**Results:** Fifty-five genes were identified as monogenic causes of nephrolithiasis. All GAPs comprised at least one extra-renal phenotype category. The median phenotype category was part of 10 (18%) unique GAPs. A total of 6 significantly overrepresented (growth, skeletal, neurologic, abdomen/gastrointestinal, muscle, metabolic features) and 5 significantly underrepresented (mouth, voice, neck, immunology, neoplasia) phenotype categories were identified among our group of monogenic kidney stone diseases (p<0.05) with impaired growth being the most common manifestation.

**Conclusion:** Monogenic nephrolithiasis is a multi-system disorder. Recognizing the extra-renal manifestations associated with monogenic causes of kidney stones is critical for earlier diagnosis and optimal prognosis in patients.

## Introduction

Nephrolithiasis is a complex disease process caused by metabolic abnormalities resulting in abnormal urinary solute concentrations and subsequent calculi formation.^1^ Epidemiological studies demonstrated increasing incidence and prevalence of kidney stones over the last few decades with kidney stones now affecting 1 in 11 people in the United States.^2,3^ This upward trend was particularly prominent in the pediatric population— the yearly incidence of kidney stones in children under the age of 18 increased from 7.2 per 100,000 children between 1984 and 1990 to a surprising 65.2 cases per 100,000 children in 2011.^4,5^

The etiology of nephrolithiasis involves a heterogenous combination of environmental, dietary, genetic, and metabolic factors.^6^ Genetics significantly contribute to kidney stone predisposition, with a positive family history in 35-60% of stone formers and a heritability of 45-50%.^7,8^ Around 12% of adults with kidney stone disease have pathogenic mutations in single genes.^9^ This monogenic causation is even more significant in the pediatric population, in which monogenic mutations have been identified in 21-29% of kidney stone cases.^10,11^ Nonetheless, given the hereditary predisposition seen in nephrolithiasis and our growing knowledge of its various genetic etiologies, the prevalence of monogenic kidney stone diseases is likely underestimated.

There has been an increasing amount of evidence linking kidney stones with the development of chronic conditions such as chronic kidney disease, bone disorders, and atherosclerosis.^12–14^Current guidelines often lack recommendations for evaluating patients with kidney stones for extra-renal comorbidities. This may result in the delayed diagnosis of comorbidities, poor preventive efforts, and worse clinical outcomes. Monogenic kidney stone diseases are prevalent among children, hence understanding their extra-renal manifestations can lead to a more comprehensive approach to evaluating this population. In this paper, we aim to explore the range of extra-renal pathologies associated with monogenic kidney stone diseases. We also highlight the organ systems and structures that are most and least commonly affected in this group.

## Methods

This research study was conducted using data obtained from public databases. Given the de-identified nature of the data, our institutional review board determined that ethics approval was not required for this study.

A literature review was performed to identify genes that are well-recognized monogenic causes of kidney stones. Diseases associated with each gene and their properties were identified using the Online Mendelian Inheritance in Man (OMIM) database, a comprehensive and curated primary repository of information on human genes, phenotypes, and their relationships, supplemental table 1.^15^ Then, OMIM clinical synopses (https://www.omim.org/search/advanced/clinicalSynopsis) were used to identify the clinical phenotypes associated with each one of these diseases. OMIM clinical synopses categorize the phenotypic features of each disease into distinct categories based on anatomical classification.^15^ We followed the same phenotypic classification system used in OMIM clinical synopses with the exception of two categories, genitourinary and laboratory abnormalities, leaving a total of 24 phenotype categories. Genitourinary and lab abnormalities were omitted because our study’s primary focus was on assessing extra-renal manifestations. Genes whose disease phenotypes were not evaluated by OMIM were excluded.

Additionally, the Human Protein Atlas (HPA) database (https://www.proteinatlas.org/) was used to identify the proteins encoded by every gene on our list, supplemental table 1.

In order to minimize the heterogeneity and inaccuracies arising from overlapping phenotypes associated with the same gene, all disease phenotypes caused by the same gene were merged into a single gene-associated phenotype (GAP) unit such that one GAP encompasses all the related disease phenotypes caused by a particular gene. The GAPs of all the given genes are summarized in supplemental table 2. The number of GAPs that are associated with each phenotype category was calculated, and the median phenotype category was determined. Subsequently, every phenotype category was classified as either overrepresented or underrepresented if the number of GAPs involving them was higher or lower than the median, respectively.

To determine if each phenotype category is significantly overrepresented or underrepresented, a chi-square test was performed comparing the median phenotype category with the number of GAPs involving each given category. In addition, we measured how many phenotypic categories each GAP involved (Supplemental Table 2). The summary of our methods is outlined in figure 1.

**Figure 1.**
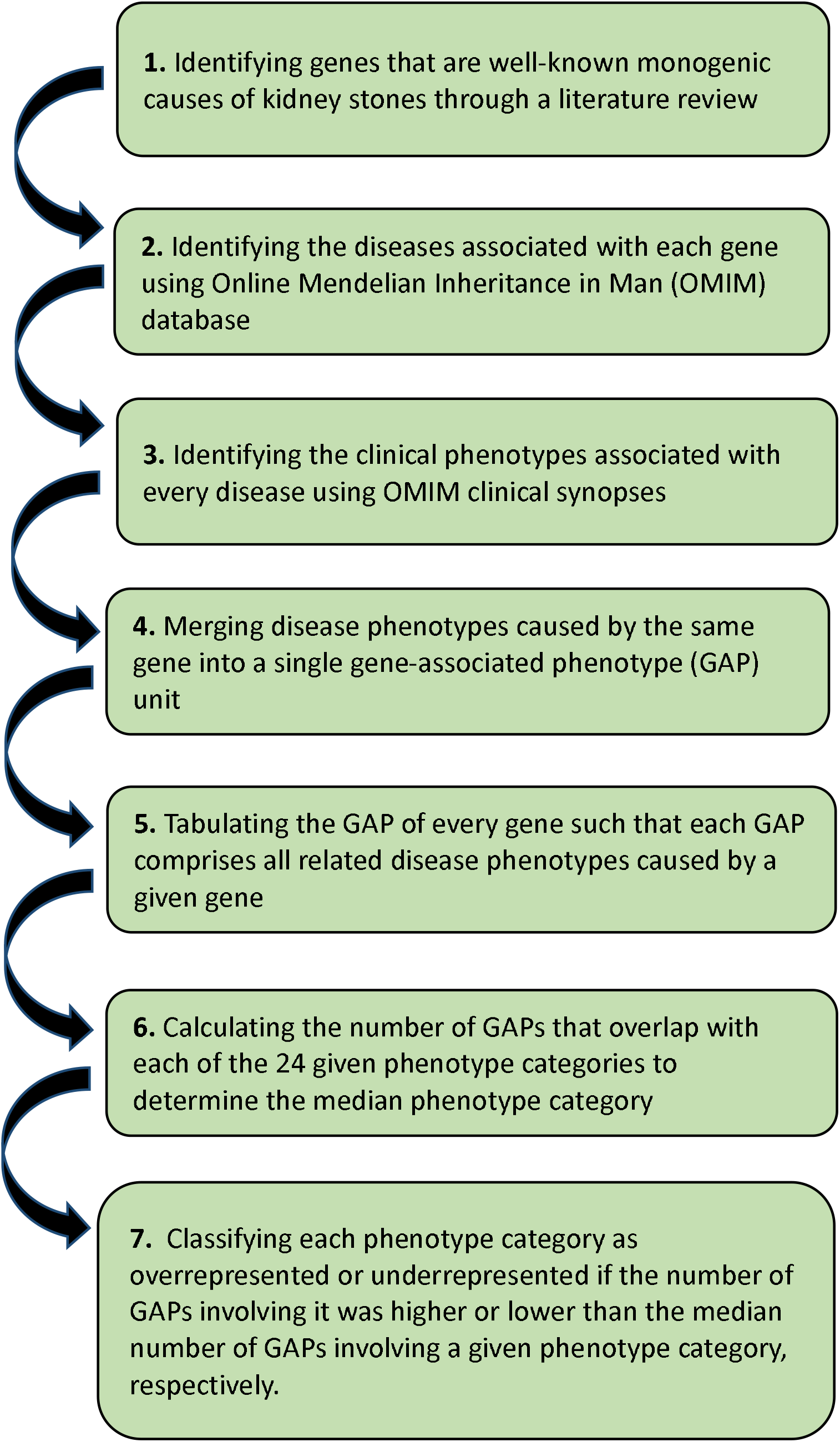
Flow diagram summarizing our methodology approach for the study

## Results

Fifty-five genes were identified to cause human nephrolithiasis as monogenic traits, supplemental table 1. These genes have been associated with 74 related diseases.

Supplemental table 1 provides a summary of the 55 genes, their encoded proteins, associated disease, and corresponding mode of inheritance. Most encoded proteins are renal solute transporters, channels, receptors, and metabolic enzymes that play a major role in the pathophysiology of monogenic kidney stone diseases. The clinical manifestations of a select group of these conditions are described in detail in Table 1. Symptom onset occurs before age 18 for the majority of these conditions. All conditions are associated with nephrolithiasis or nephrocalcinosis, as well as extra-renal manifestations. These additional symptoms include impaired growth and skeletal development, neurological deficits, gastrointestinal disruptions, hormonal imbalances, cardiovascular conditions, as well as various structural and functional abnormalities, as outlined in Table 1.

**Table 1.**
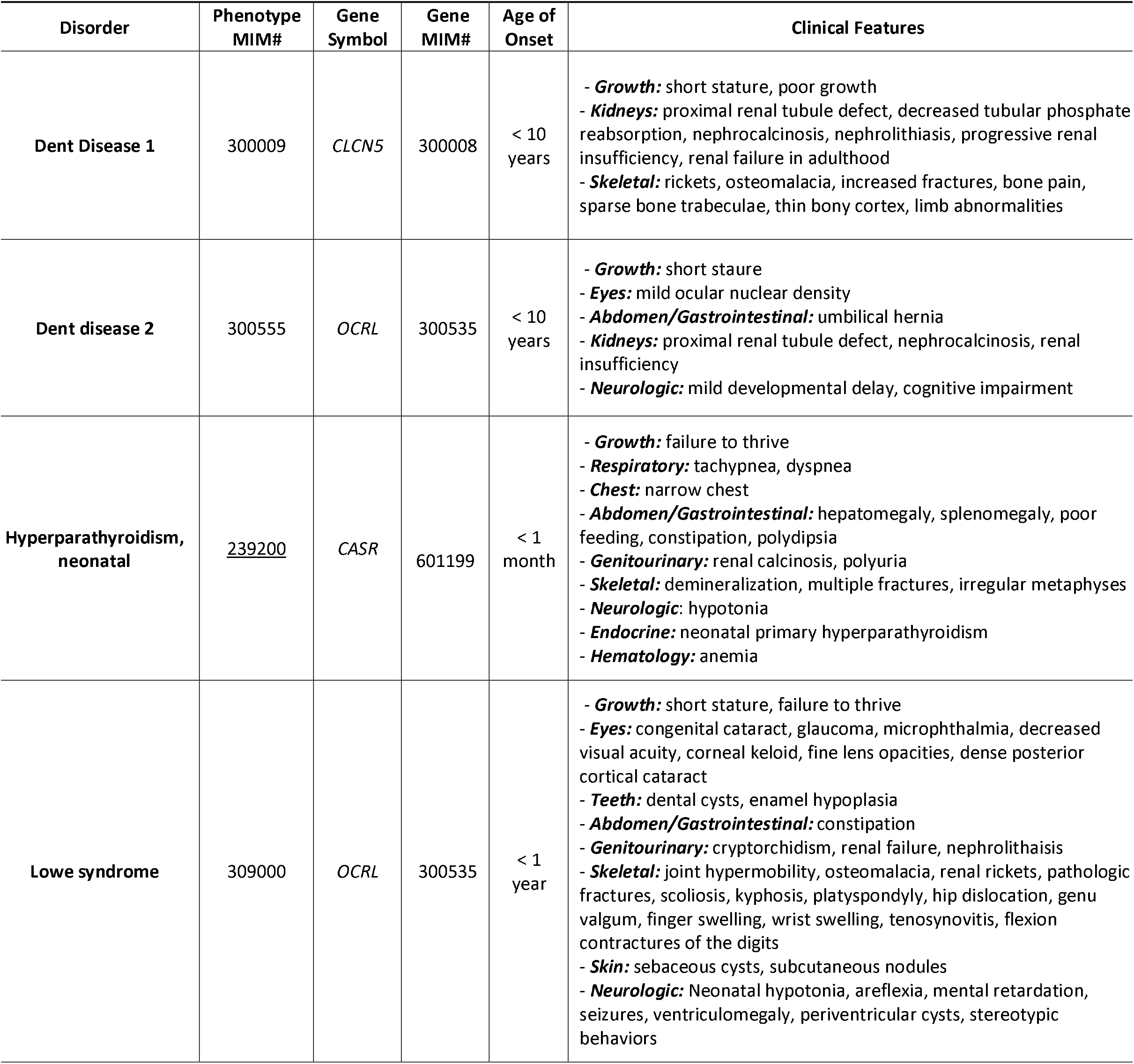
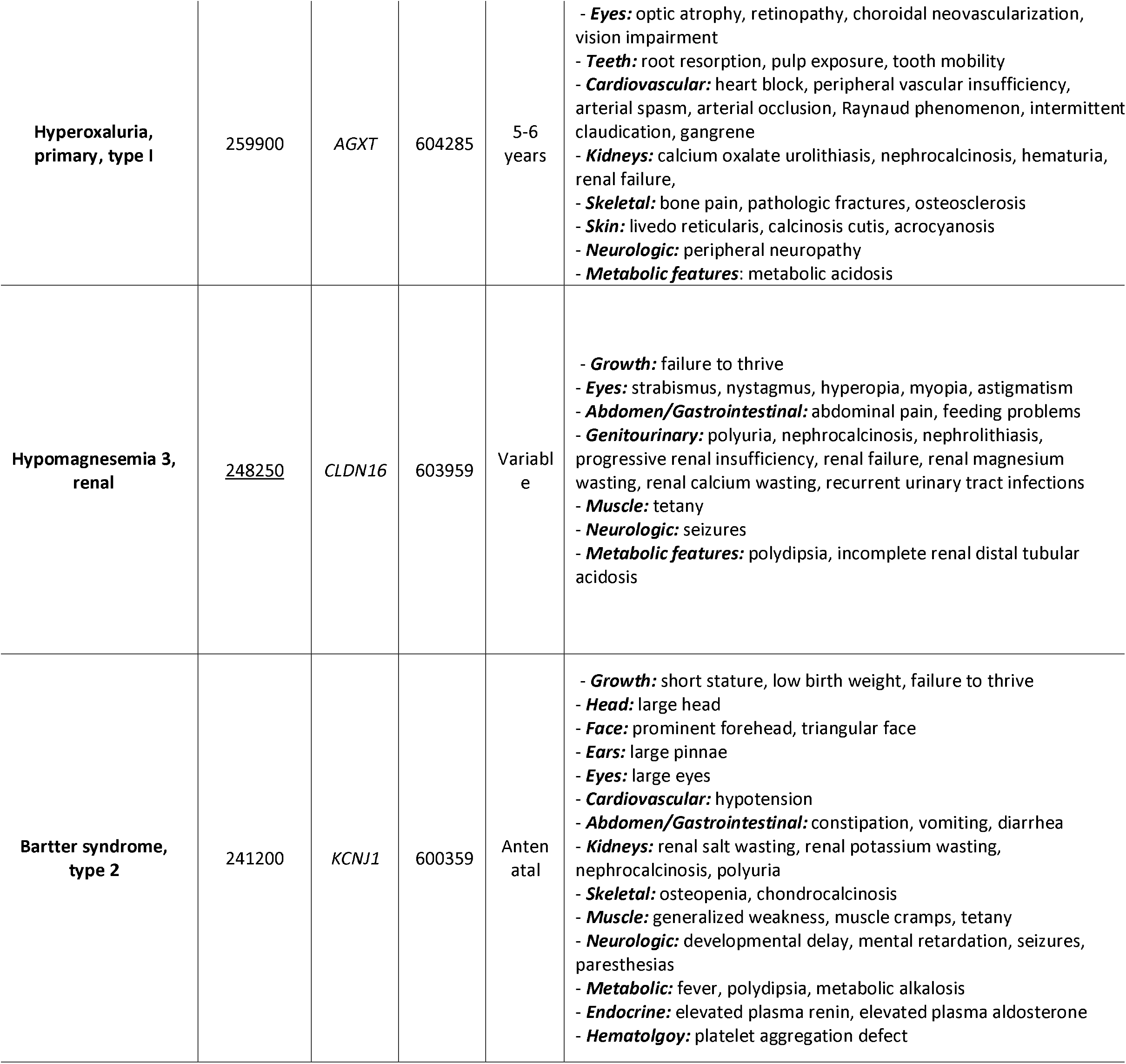
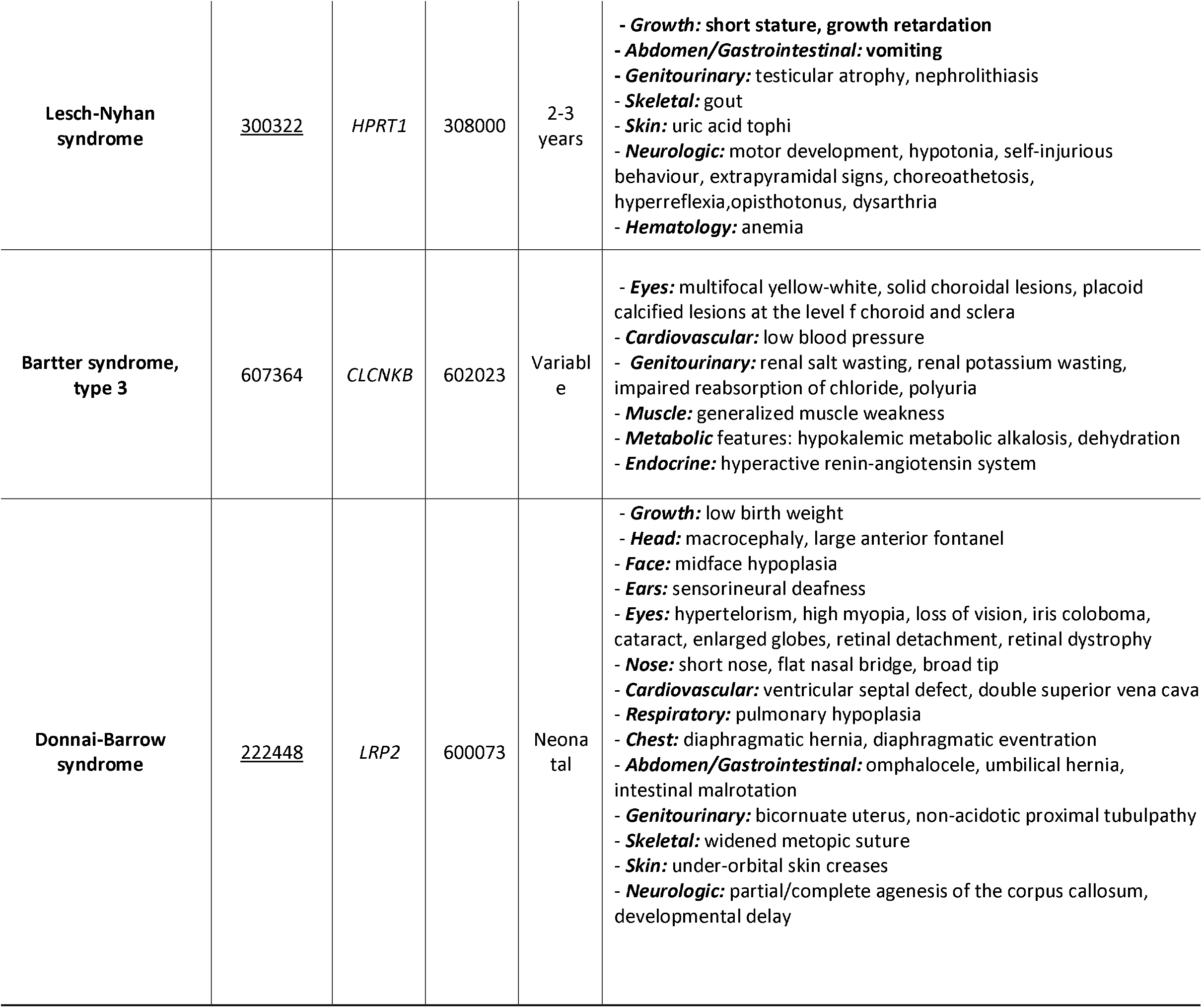
Description of selected monogenic kidney stone diseases showing their OMIM number, gene, age of onset, and clinical features.

Figure 2 shows the percentage of GAPs involving each of the 24 different phenotype categories listed in OMIM. The median phenotype category was part of 10 GAPs (18%). There was a total of 6 significantly overrepresented and 5 significantly underrepresented phenotype categories. Among our list of 55 GAPs, thirty-one (56%) GAPs were found to impact growth, making it the most affected non-renal phenotype in patients with monogenic kidney stone diseases (Figure 2, Supplemental Table 2). Pathologies affecting the growth (56%, n=31), skeletal system (53%, n=29), neurologic system (53%, n=29), abdomen/gastrointestinal system (45%, n=25), muscles (40%, n=22), and metabolic status (36%, n=20) were also significantly overrepresented in a large group of monogenic kidney stone diseases (all *P*<0.05), Figure 2 and Supplemental Table 2. On the other hand, abnormalities of the mouth (5%, n=3), voice (5%, n=3), neck (2%, n=1), immunological disorders (2%, n=1), and neoplastic disorders (2%, n=1) are hardly reported in monogenic kidney stone diseases and significantly underrepresented in this population (all *P*<0.05) (Figure 2 and Supplemental Table 2). Other phenotypic categories were variably affected in monogenic kidney stone diseases, and were not significantly overrepresented in this population(Figure 2 and Supplemental Table 2).

**Figure 2.**
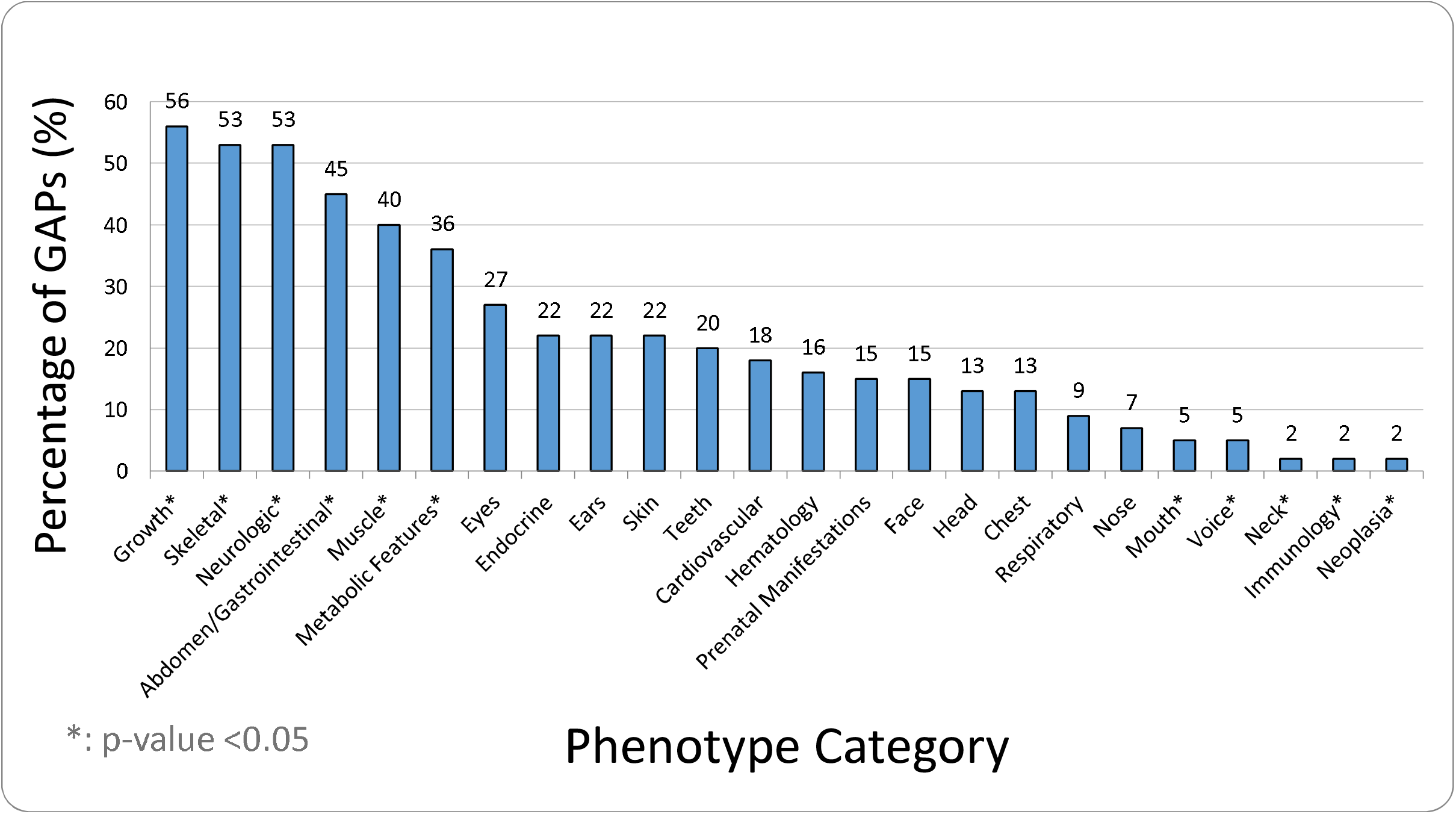
A histogram showing the percentage of genes (%) affecting each of the 24 different phenotype categories listed in OMIM. With the median percentage of genes affecting a given phenotype category being 18% (n=10), any category with a percentage of genes affecting them higher than 18% was considered overrepresented, and those with a percentage of genes affecting them lower than 18% were considered underrepresented. Using chi-square test, our analysis revealed that only growth, skeletal, neurologic, abdomen, muscle, and metabolic features were significantly overrepresented (truly deviated from the median), and only mouth, voice, neck, immunology, and neoplasia were significantly underrepresented among our list of genes (p<0.05). GAP: gene-associated phenotype

Figure 3 shows the percentage of GAPs affecting predefined ranges of numbers of phenotype categories. None of the 55 GAPs were associated with isolated nephrolithiasis or nephrocalcinosis alone; they all affected at least one phenotype category outside the kidneys. Specifically, 36% (n=20) of GAPs impacted 1-3 categories, 16% (n=9) impacted 4-6 categories, and 20% (n=11) impacted 7-9 categories. Interestingly, 27% (n=15) of the listed GAPs affected at least 10 categories. The number of phenotype categories comprising every GAP is highlighted in supplemental table 2.

**Figure 3.**
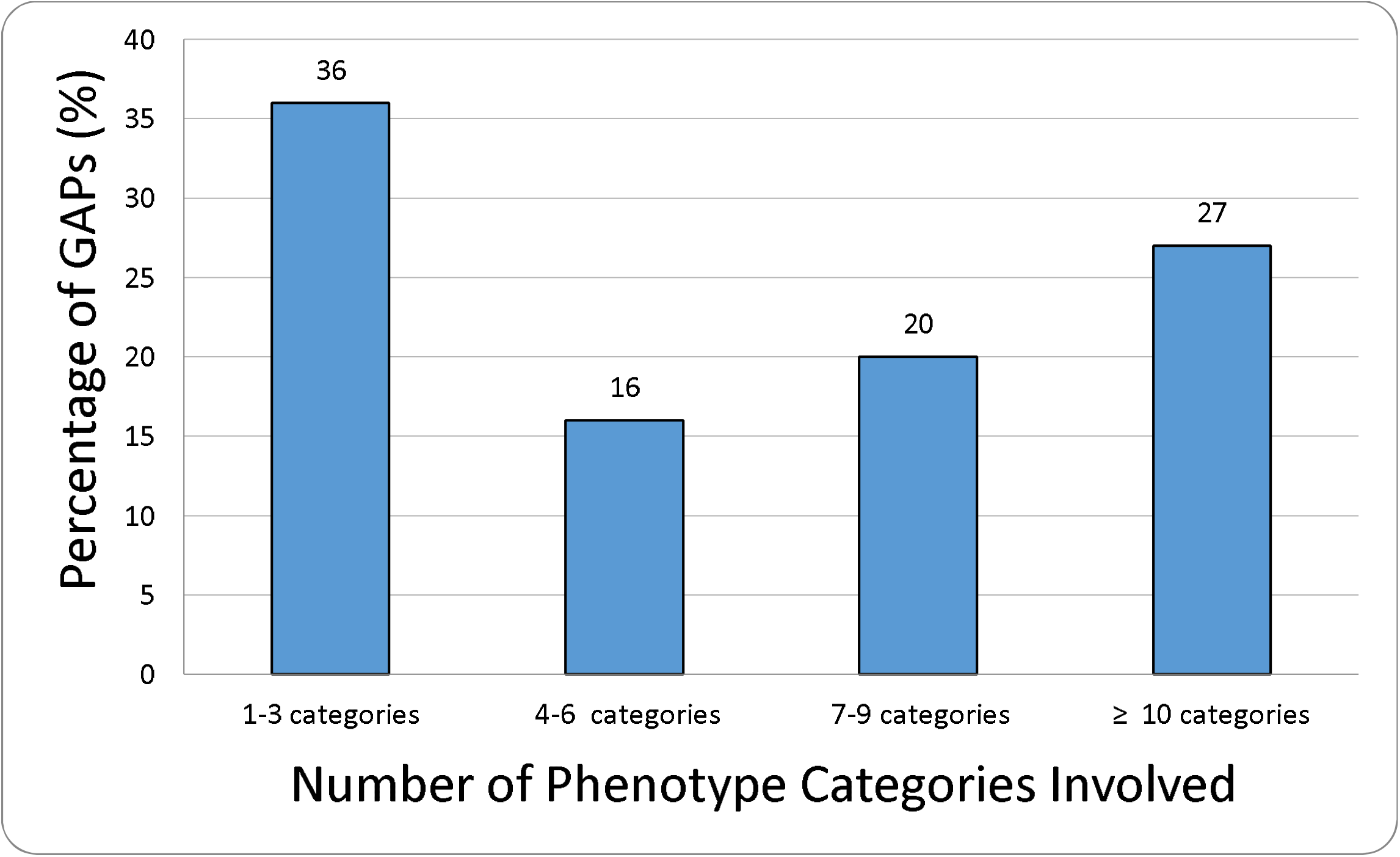
A histogram showing the percentage of genes affecting predefined ranges of number of phenotype categories. GAP: gene-associated phenotype

## Discussion

Identifying individuals with underlying monogenic causes of nephrolithiasis can be a challenging task. Despite its increased availability and recent advancements, genetic testing is still not a first-line diagnostic tool in patients with kidney stone disease.^16,17^Hence, a high index of clinical suspicion is warranted to pursue genetic testing.^18^Certain aspects of stone-formers’ history and presentation may hint towards a genetic etiology and should prompt a comprehensive investigation in order to ensure optimal treatment and prognosis. Pertinent history may include an early onset of stone disease, strong family history, high recurrence rate, bilateral/multiple stones, associated nephrocalcinosis, renal failure, and extra-renal manifestations.^19–21^ It is crucial for treating physicians to be aware of the extra-renal complications and risks associated with the disease.

### Growth and Skeletal Abnormalities

Our findings reveal that impaired growth and skeletal abnormalities such as short stature, rickets, and osteoporosis, are significantly overrepresented in patients with monogenic kidney stone diseases.

Many studies evaluating bone disease in stone formers have found low bone mineral density (BMD) and a higher risk of vertebral fractures.^12,12^According to a study by Jaeger et al., stone formers were slightly shorter and had a significantly lower BMD in the tibial epiphysis and diaphysis compared to the control group.^22^ Evidence has also shown increased markers of bone turnover in this population.^23^ Even though the mechanism of bone loss in stone formers is not fully understood, it can be partly attributed to the hypercalciuric state seen in most patients with calcium-based kidney stones.^24^ It is possible that the renal wasting of calcium and its net loss from the body can contribute to low BMD and weaker bone structure.^25^ For example, hypercalciuria and hypophosphatemia are common features in Dent Disease 1 and hypophosphatemic rickets with hypercalciuria, which are caused by pathogenic variants in *CLCN5* and *SLC34A3*, respectively supplemental table 1. Both of these conditions are associated with rickets, osteomalacia, bone pain, and increased rate of fractures.^26^ Because childhood and adolescence are critical periods for attaining bone mass, it is crucial to identify children at risk for low BMD to prevent impairment of overall growth and skeletal development.^12^

### Neurological Abnormalities

Our study shows that neurologic impairments are significantly overrepresented in individuals with monogenic kidney stone diseases. Increasing awareness of these conditions among physicians may aid in early diagnosis of neurologic involvement and initiate subsequent care or monitoring. These manifestations result from common pathophysiological processes that simultaneously affect the nervous system and the kidneys.^27^For example, Lowe syndrome, caused by pathogenic variants in the *OCRL* gene, is a multisystemic disease associated with ocular, renal, and neurologic manifestations.^28^ Loss of OCRL function disrupts important cellular processes like endocytic trafficking, leading to cataracts, renal tubular dysfunction, hypotonia, areflexia, seizures, and behavioral abnormalities.^29^ Pathogenic variants in *KCNJ10*, which encodes for an inwardly rectifying potassium channel, causes accumulation of potassium in the extracellular space and leads to epilepsy, ataxia, sensorineural deafness, and tubulopathy (EAST syndrome).^30^

### Gastrointestinal, Muscular, and Metabolic Manifestations

The majority of monogenic kidney stone diseases are characterized by renal tubular or glomerular dysfunction, leading to various metabolic and electrolyte abnormalities.^31^These physiologic derangements can contribute to a wide range of gastrointestinal and muscular issues, which we found to be significantly overrepresented in monogenic stone diseases.^27^ For instance, pathogenic variants in the *CASR* gene can cause defective calcium sensing receptors, resulting in hypocalcemia and subsequent muscle spasms and tetany.^32^Similarly, pathogenic variants in the *ALDOB* gene can cause hereditary fructose intolerance, which is often associated with hypophosphatemia, metabolic acidosis, and subsequent symptoms such as vomiting, nausea, malnutrition, and abdominal pain.^33^

All GAPs included in our study were found to impact at least one phenotype category beyond the urinary system, indicating their influence on various bodily functions and the depth of their involvement. Interestingly, despite being present in some monogenic kidney stone diseases, immunological and neoplastic disorders were underrepresented in this group. This suggests that patients with genetic kidney stone disease are less likely to develop these conditions. Additionally, we found that monogenic kidney stone diseases can affect other systems, such as the cardiovascular, endocrine, and hematological system, which is consistent with previous studies.^14,21,34^

The management of nephrolithiasis has made significant strides in recent decades, with a focus on secondary prevention through dietary and lifestyle recommendations for adults.^35,36^ However, current guidelines for managing pediatric stone patients are inadequate, leaving many monogenic kidney stone diseases, which often manifest at a young age and involve multiple systems, underdiagnosed and undertreated.^37^ This calls for a more comprehensive approach that goes beyond the urinary system and considers the broader spectrum of associated pathologies. By leveraging existing knowledge of relevant extra-renal pathologies, healthcare providers can improve the diagnosis and management of genetic kidney stone diseases in patients of all ages.

### Limitations

This study is limited by what is currently known about the genetic etiologies of nephrolithiasis. There were 55 genes identified, along with their associated pathologies, which may not represent the complete picture of kidney stone genetics. As our understanding of the genetic basis of nephrolithiasis evolves, additional genes and extra-renal pathologies may come to light, which could alter our findings. Therefore, these research findings are dynamic and subject to change with time.

Furthermore, this study focused on individuals with monogenic kidney stone diseases but not general stone formers. Hence, the generalizability of our results and the extent to which they overlap with other stone formers can only be ascertained with future research.

## Conclusion

Our work highlights the most critical extra-renal pathologies in patients with genetic kidney stone diseases. With the incidence of pediatric kidney stones on the rise, urologists will encounter a growing number of patients who developed nephrolithiasis during childhood, many of whom may have underlying monogenic stone disease.^38^ As a result, it is crucial for treating physicians to not only manage the acute episodes of kidney stones, but also be aware of the extra-renal complications and risks associated.

## Data Availability

All data produced in the present study are available upon reasonable request to the authors

## Acknowledgements

None

## Ethics approval statement

### Research involving Human Participants and/or Animals

This is an observational study. The data used is de-identified and publicly available. The Case Western Reserve University/University Hospital institutional review board has confirmed that no ethical approval is required.

### Informed consent

Given the deidentified nature of the data, the Case Western Reserve University/University Hospital institutional review board determined that this observational study did not constitute human participants research and was thus exempted from review and the need for informed consent, in accordance with 45 CFR §46.

## Author Contribution

All listed authors contributed to this manuscript through data collection, analysis, interpretation, writing, and reviewing the manuscript.

## References

1. Alelign T, Petros B. Kidney Stone Disease: An Update on Current Concepts. Adv Urol. 2018;2018:3068365. doi:10.1155/2018/3068365

2. Scales CD, Smith AC, Hanley JM, Saigal CS. Prevalence of Kidney Stones in the United States. Eur Urol. 2012;62(1):160–165. doi:10.1016/j.eururo.2012.03.052

3. Tundo G, Vollstedt A, Meeks W, Pais V. Beyond Prevalence: Annual Cumulative Incidence of Kidney Stones in the United States. Journal of Urology. 2021;205(6):1704–1709. doi:10.1097/JU.0000000000001629

4. Ward JB, Feinstein L, Pierce C, et al. Pediatric Urinary Stone Disease in the United States: The Urologic Diseases in America Project. Urology. 2019;129:180–187. doi:10.1016/j.urology.2019.04.012

5. Dwyer ME, Krambeck AE, Bergstralh EJ, Milliner DS, Lieske JC, Rule AD. Temporal trends in incidence of kidney stones among children: a 25-year population based study. J Urol. 2012;188(1):247–252. doi:10.1016/j.juro.2012.03.021

6. Moe OW. Kidney stones: pathophysiology and medical management. Lancet. 2006;367(9507):333–344. doi:10.1016/S0140-6736(06)68071-9

7. Goldfarb DS, Avery AR, Beara-Lasic L, Duncan GE, Goldberg J. A Twin Study of Genetic Influences on Nephrolithiasis in Women and Men. Kidney Int Rep. 2018;4(4):535–540. doi:10.1016/j.ekir.2018.11.017

8. Lieske JC, Wang X. Heritable traits that contribute to nephrolithiasis. Urolithiasis. 2019;47(1):5–10. doi:10.1007/s00240-018-1095-1

9. Howles SA, Thakker RV. Genetics of kidney stone disease. Nat Rev Urol. 2020;17(7):407–421. doi:10.1038/s41585-020-0332-x

10. Daga A, Majmundar AJ, Braun DA, et al. Whole exome sequencing frequently detects a monogenic cause in early onset nephrolithiasis and nephrocalcinosis. Kidney Int. 2018;93(1):204–213. doi:10.1016/j.kint.2017.06.025

11. Huang L, Qi C, Zhu G, et al. Genetic testing enables a precision medicine approach for nephrolithiasis and nephrocalcinosis in pediatrics: a single-center cohort. Mol Genet Genomics. 2022;297(4):1049–1061. doi:10.1007/s00438-022-01897-z

12. Schwaderer AL, Kusumi K, Ayoob RM. Pediatric nephrolithiasis and the link to bone metabolism. Curr Opin Pediatr. 2014;26(2):207–214. doi:10.1097/MOP.0000000000000069

13. Ferraro PM, Bargagli M, Trinchieri A, Gambaro G. Risk of Kidney Stones: Influence of Dietary Factors, Dietary Patterns, and Vegetarian-Vegan Diets. Nutrients. 2020;12(3):E779. doi:10.3390/nu12030779

14. Alexander RT, Hemmelgarn BR, Wiebe N, et al. Kidney stones and kidney function loss: a cohort study. BMJ. 2012;345:e5287. doi:10.1136/bmj.e5287

15. Amberger JS, Bocchini CA, Schiettecatte F, Scott AF, Hamosh A. OMIM.org: Online Mendelian Inheritance in Man (OMIM®), an online catalog of human genes and genetic disorders. Nucleic Acids Res. 2015;43(Database issue):D789–798. doi:10.1093/nar/gku1205

16. Vasudevan V, Samson P, Smith AD, Okeke Z. The genetic framework for development of nephrolithiasis. Asian J Urol. 2017;4(1):18–26. doi:10.1016/j.ajur.2016.11.003

17. Bhojani N, Bjazevic J, Wallace B, et al. UPDATE – Canadian Urological Association guideline: Evaluation and medical management of kidney stones. Can Urol Assoc J. 2022;16(6):175–188. doi:10.5489/cuaj.7872

18. Maalouf NM, Tondapu P, Guth ES, Livingston EH, Sakhaee K. Hypocitraturia and hyperoxaluria after Roux-en-Y gastric bypass surgery. J Urol. 2010;183(3):1026–1030. doi:10.1016/j.juro.2009.11.022

19. Curhan GC, Willett WC, Rimm EB, Stampfer MJ. Family history and risk of kidney stones. J Am Soc Nephrol. 1997;8(10):1568–1573. doi:10.1681/ASN.V8101568

20. Jungers P, Joly D, Blanchard A, Courbebaisse M, Knebelmann B, Daudon M. [Inherited monogenic kidney stone diseases: recent diagnostic and therapeutic advances]. Nephrol Ther. 2008;4(4):231–255. doi:10.1016/j.nephro.2007.12.005

21. Ferraro PM, Taylor EN, Eisner BH, et al. History of kidney stones and the risk of coronary heart disease. JAMA. 2013;310(4):408–415. doi:10.1001/jama.2013.8780

22. Jaeger P, Lippuner K, Casez JP, Hess B, Ackermann D, Hug C. Low bone mass in idiopathic renal stone formers: magnitude and significance. J Bone Miner Res. 1994;9(10):1525–1532. doi:10.1002/jbmr.5650091004

23. Krieger NS, Bushinsky DA. The Relation between Bone and Stone Formation. Calcif Tissue Int. 2013;93(4):374–381. doi:10.1007/s00223-012-9686-2

24. Pietschmann F, Breslau NA, Pak CY. Reduced vertebral bone density in hypercalciuric nephrolithiasis. J Bone Miner Res. 1992;7(12):1383–1388. doi:10.1002/jbmr.5650071205

25. Sella S, Cattelan C, Realdi G, Giannini S. Bone disease in primary hypercalciuria. Clin Cases Miner Bone Metab. 2008;5(2):118–126.

26. Schott C, Pourtousi A, Connaughton DM. Monogenic causation of pediatric nephrolithiasis. Frontiers in Urology. 2022;2. Accessed January 18, 2023. https://www.frontiersin.org/articles/10.3389/fruro.2022.1075711

27. Bhowmick SS, Lang AE. Movement Disorders and Renal Diseases. Mov Disord Clin Pract. 2020;7(7):763–779. doi:10.1002/mdc3.13005

28. Attree O, Olivos IM, Okabe I, et al. The Lowe’s oculocerebrorenal syndrome gene encodes a protein highly homologous to inositol polyphosphate-5-phosphatase. Nature. 1992;358(6383):239–242. doi:10.1038/358239a0

29. Romani M, Micalizzi A, Valente EM. Joubert syndrome: congenital cerebellar ataxia with the molar tooth. Lancet Neurol. 2013;12(9):894–905. doi:10.1016/S1474-4422(13)70136-4

30. Bockenhauer D, Feather S, Stanescu HC, et al. Epilepsy, ataxia, sensorineural deafness, tubulopathy, and KCNJ10 mutations. N Engl J Med. 2009;360(19):1960–1970. doi:10.1056/NEJMoa0810276

31. Rumsby G. Genetic defects underlying renal stone disease. International Journal of Surgery. 2016;36:590–595. doi:10.1016/j.ijsu.2016.11.015

32. Hendy GN, D’Souza-Li L, Yang B, Canaff L, Cole DE. Mutations of the calcium-sensing receptor (CASR) in familial hypocalciuric hypercalcemia, neonatal severe hyperparathyroidism, and autosomal dominant hypocalcemia. Hum Mutat. 2000;16(4):281–296. doi:10.1002/1098-1004(200010)16:4<281::AID-HUMU1>3.0.CO;2-A

33. Paolella G, Santamaria R, Buono P, Salvatore F. Mapping of a restriction fragment length polymorphism within the human aldolase B gene. Hum Genet. 1987;77(2):115–117. doi:10.1007/BF00272375

34. Madore F, Stampfer MJ, Rimm EB, Curhan GC. Nephrolithiasis and risk of hypertension. Am J Hypertens. 1998;11(1 Pt 1):46–53. doi:10.1016/s0895-7061(97)00371-3

35. Assimos D, Krambeck A, Miller NL, et al. Surgical Management of Stones: American Urological Association/Endourological Society Guideline, PART I. J Urol. 2016;196(4):1153–1160. doi:10.1016/j.juro.2016.05.090

36. Skolarikos A, Straub M, Knoll T, et al. Metabolic evaluation and recurrence prevention for urinary stone patients: EAU guidelines. Eur Urol. 2015;67(4):750–763. doi:10.1016/j.eururo.2014.10.029

37. Bonzo JR, Tasian GE. The Emergence of Kidney Stone Disease During Childhood-Impact on Adults. Curr Urol Rep. 2017;18(6):44. doi:10.1007/s11934-017-0691-x

38. Sas DJ, Enders FT, Mehta RA, et al. Clinical features of genetically confirmed patients with primary hyperoxaluria identified by clinical indication versus familial screening. Kidney Int. 2020;97(4):786–792. doi:10.1016/j.kint.2019.11.023

