## supplementary table 1 for "Beyond the Kidney: Extra-Renal Manifestations of Monogenic Nephrolithiasis and Their Significance"

| *Gene Symbol (n=55)* | Gene MIM # | Protein | Related Disease | Phenotype MIM # | Mode of Inheritance |
|---|---|---|---|---|---|
| ***ADCY10***[1] | 605205 | Adenylate cyclase 10 | **Hypercalciuria, absorptive, susceptibility to** | 143870 | AD |
| ***AGXT***[2] | 604285 | Alanine--glyoxylate and serine--pyruvate aminotransferase | **Hyperoxaluria, primary, type I** | 259900 | AR |
| ***ALDOB***[3] | 612724 | Aldolase, fructose-bisphosphate B | **Fructose intolerance, hereditary** | 229600 | AR |
| ***ALPL***[4] | 171760 | Alkaline phosphatase | **Hypophosphotasia, adult** | 146300 | AD, AR |
| | | | **Hypophosphotasia, child** | 241510 | AR |
| | | | **Hypophosphotasia infantile** | 241500 | AR |
| ***APRT***[5] | 102600 | Adenine phosphoribosyltransferase | **Adenine phosphoribosyltransferase deficiency, Urolithiasis (DHA stones), renal failure** | 614723 | AR |
| ***ATP6V0A4***[6] | 605239 | ATPase H+ transporting V0 subunit a4 | **Distal renal tubular acidosis 3, with or without progressive sensorineural hearing loss** | 602722 | AR |
| ***ATP6V1B1***[7] | 192132 | ATPase H+ transporting V1 subunit B1 | **Distal renal tubular acidosis 2 with progressive sensorineural hearing loss** | 267300 | AR |
| ***ATP7B***[8] | 606882 | ATPase copper transporting beta | **Wilson Disease** | 277900 | AR |
| ***BSND***[9] | 606412 | Barttin CLCNK type accessory subunit beta | **Bartter syndrome, type 4a, neonatal, with sensorineural deafness** | 602522 | AR |
| ***CA2***[10] | 611492 | Carbonic anhydrase 2 | **Osteopetrosis, autosomal recessive 3, with renal tubular acidosis** | 259730 | AR |
| ***CASR***[11] | 601199 | Calcium sensing receptor | **Hyperparathyroidism, neonatal** | 239200 | AD, AR |
| | | | **Hypocalcemia, autosomal dominant** | 601198 | AD |
| | | | **Hypocalcemia, autosomal dominant, with Bartter syndrome** | 601198 | AD |
| | | | **Hypocalciuric hypercalcemia, type I** | 145980 | AD |
| | | | **Epilepsy idiopathic generalized, susceptibility to, 8** | 612899 | N/A |
| ***CLCN5***[12] | 300008 | Chloride voltage-gated channel 5 | **Dent Disease 1** | 300009 | XLR |
| | | | **Hypophosphatemic rickets** | 300554 | XLR |
| | | | **Nephrolithiasis, type I** | 310486 | XLR |
| | | | **Proteinuria, low molecular weight, with hypercalciuric nephrocalcinosis** | 308990 | XLR |
| ***CLCNKA***[13] | 602024 | Chloride voltage-gated channel Ka | **Bartter syndrome, type 4b, digenic** | 613090 | DR |
| ***CLCNKB***[14] | 602023 | Chloride voltage-gated channel Kb | **Bartter syndrome, type 4b, digenic** | 613090 | DR |

| | | | | | |
|---|---|---|---|---|---|
| | | | **Bartter syndrome, type 3** | 607364 | AR |
| ***CLDN16***[15] | 603959 | Claudin 16 | **Hypomagnesemia 3, renal** | 248250 | AR |
| ***CLDN19***[16] | 610036 | Claudin 19 | **Hypomagnesemia 5, renal, with ocular involvement** | 248190 | AR |
| ***CTNS***[17] | 606272 | Cystinosin, lysosomal cystine transporter | **Cytinosis, typical/atypical nephropathic** | 219800 | AR |
| | | | **Cystinosis, late-onset juvenile or adolesent nephropathic** | 219900 | AR |
| ***CYP24A1***[18] | 126065 | Cytochrome P450 family 24 subfamily A member 1 | **Hypercalcemia, infantile, 1** | 143880 | AR |
| ***FAH***[19] | 613871 | Fumarylacetoacetate hydrolase | **Tyrosinemia, type I** | 276700 | AR |
| ***FAM20A***[20] | 611062 | FAM20A golgi associated secretory pathway pseudokinase | **Amelogenesis imperfecta, type IG (enamel-renal syndrome)** | 204690 | AR |
| ***FOXI1***[21] | 601093 | Forkhead box I1 | **Enlarged vestibular aqueduct** | 600791 | AR |
| ***G6PC***[22] | 613742 | Glucose-6-phosphatase catalytic subunit | **Glycogen storage disease Ia** | 232200 | AR |
| ***GNA11***[23] | 139313 | G protein subunit alpha 11 | **Hypocalcemia, autosomal dominant 2** | 615361 | AD |
| | | | **Hypocalciuric hypercalcemia, type II** | 145981 | AD |
| ***GPHN***[24] | 603930 | Gephyrin | **Molybdenum cofactor deficiency C** | 615501 | AR |
| ***GRHPR***[25] | 604269 | Glyoxylate and hydroxypyruvate reductase | **Hyperoxaluria, primary, type II** | 260000 | AR |
| ***HNF4A***[26] | 600281 | Hepatocyte nuclear factor 4 alpha | **Fanconi renotubular syndrome 4, with maturity-onset diabetes of the young** | 616026 | AD |
| ***HOGA1***[27] | 613597 | 4-hydroxy-2-oxoglutarate aldolase 1 | **Hyperoxaluria, primary, type III** | 613616 | AR |
| ***HPRT1***[28] | 308000 | Hypoxanthine phosphoribosyltransferase 1 | **Kelley-Seegmiller syndrome, partial HPRT defiency, HPRT related gout** | 300323 | XLR |
| | | | **Lesch-Nyhan syndrome** | 300322 | XLR |
| ***KCNJ1***[29] | 600359 | Potassium inwardly rectifying channel subfamily J member 1 | **Bartter syndrome, type 2** | 241200 | AR |
| ***KCNJ10***[30] | 602208 | Potassium inwardly rectifying channel subfamily J member 10 | **SESAME syndrome** | 612780 | AR |
| ***KL***[31] | 604824 | Klotho | **Tumoral calcinosis, hyperphosphatemic, familial, 3** | 617994 | AR |
| ***LRP2***[32] | 600073 | LDL receptor related protein 2 | **Donnai-Barrow syndrome** | 222448 | AR |
| ***MAGED2***[33] | 300470 | MAGE family member D2 | **Bartter syndrome, type 5, antenatal, transient** | 300971 | XLR |
| ***MGP***[34] | 154870 | Matrix Gla protein | **Keutel syndrome** | 245150 | AR |
| ***MOCOS***[35] | 613274 | Molybdenum cofactor sulfurase | **Xanthinuria, type II** | 603592 | AR |

| | | | | | |
|---|---|---|---|---|---|
| ***MOCS1***[36] | 603707 | Molybdenum cofactor synthesis 1 | **Molybdenum cofactor deficiency A** | 252150 | AR |
| ***MOCS2***[36] | 603708 | Molybdenum cofactor synthesis 2 | **Molybdenum cofactor deficiency B** | 252160 | AR |
| ***OCRL***[37] | 300535 | OCRL inositol polyphosphate-5-phosphatase | **Dent disease 2** | 300555 | XLR |
| | | | **Lowe syndrome** | 309000 | XLR |
| ***ORAI1***[38] | 610277 | ORAI calcium release-activated calcium modulator 1 | **Immunodefiency 9** | 612782 | AR |
| | | | **Myopathy, tubular aggregate, 2** | 615883 | AD |
| ***PRPS1***[39] | 311850 | Phosphoribosyl pyrophosphate synthetase 1 | **Phosphoribosylpyrophosphate synthetase superactivity** | 300661 | XLR |
| ***SLC12A1***[29] | 600839 | Solute carrier family 12 member 1 | **Bartter syndrome, type 1** | 601678 | AR |
| ***SLC22A12***[40] | 607096 | Solute carrier family 22 member 12 | **Hypouricemia, renal (RHUC1)** | 220150 | AR, AD |
| ***SLC26A1***[41] | 610130 | Solute carrier family 26 member 1 | **Nephrolithiasis, calcium oxalate** | 167030 | AR |
| ***SLC2A2***[42] | 138160 | Solute carrier family 2 member 2 | **Fanconi-Bickel Syndrome** | 227810 | AR |
| ***SLC2A9***[43] | 606142 | Solute carrier family 2 member 9 | **Renal hypouricemia, RHUC2** | 612076 | AR, AD |
| ***SLC34A1***[44] | 182309 | Solute carrier family 34 member 1 | **Hypercalcemia, infantile, 2** | 616963 | AR |
| | | | **Nephrolithiasis/osteoporosis, hypophosphatemic, 1** | 612286 | AD |
| | | | **Fanconi renotubular syndrome 2** | 613388 | AR |
| | | | **Hypercalcemia, infantile, 2** | 616963 | AR |
| ***SLC34A3***[45] | 609826 | Solute carrier family 34 member 3 | **Hypophosphatemic rickets with hypercalciuria** | 241530 | AR |
| ***SLC3A1***[46] | 104614 | Solute carrier family 3 member 1 | **Cystinuria** | 220100 | AD, AR |
| ***SLC4A1***[47] | 109270 | Solute carrier family 4 member 1 | **Distal renal tubular acidosis 1** | 179800 | AD |
| | | | **Distal renal tubular acidosis 4 with hemolytic anemia** | 611590 | AR |
| ***SLC5A1***[48] | 82380 | Solute carrier family 5 member 1 | **Glucose/galactose malabsorption** | 606824 | AR |
| ***SLC7A9***[49] | 604144 | Solute carrier family 7 member 9 | **Cystinuria** | 220100 | AD, AR |
| ***SLC9A3R1***[50] | 604990 | SLC9A3 regulator 1 | **Hypophosphatemic nephrolithiasis/osteoporosis 2, (NPHLOP2)** | 612287 | AD |
| ***VDR***[51] | 601769 | Vitamin D receptor | **Rickets, vitamin D-resistant, type IIA; Idiopathic hypercalcuria** | 277440 | AR |
| ***WDR72***[52] | 613214 | WD repeat domain 72 | **Amelogenesis imperfecta, type IIA3** | 613211 | AR |
| ***XDH***[53] | 607633 | Xanthine dehydrogenase | **Xanthinuria, type I** | 278300 | AR |

**Supplemental Table 1. List of genes known to cause monogenic kidney stone disease along with their corresponding MIM number, protein, related disorder, and mode of inheritance.**

1. Reed BY, Gitomer WL, Heller HJ, et al. Identification and characterization of a gene with base substitutions associated with the absorptive hypercalciuria phenotype and low spinal bone density. *J Clin Endocrinol Metab*. 2002;87(4):1476-1485. doi:10.1210/jcem.87.4.8300

2. Purdue PE, Allsop J, Isaya G, Rosenberg LE, Danpure CJ. Mistargeting of peroxisomal L-alanine:glyoxylate aminotransferase to mitochondria in primary hyperoxaluria patients depends upon activation of a cryptic mitochondrial targeting sequence by a point mutation. *Proc Natl Acad Sci U S A*. 1991;88(23):10900-10904. doi:10.1073/pnas.88.23.10900

3. Paolella G, Santamaria R, Buono P, Salvatore F. Mapping of a restriction fragment length polymorphism within the human aldolase B gene. *Hum Genet*. 1987;77(2):115-117. doi:10.1007/BF00272375

4. Whyte MP, Rettinger SD, Vrabel LA. Infantile hypophosphatasia: enzymatic defect explored with alkaline phosphatase-deficient skin fibroblasts in culture. *Calcif Tissue Int*. 1987;40(5):244-252. doi:10.1007/BF02555256

5. Kelley WN, Levy RI, Rosenbloom FM, Henderson JF, Seegmiller JE. Adenine phosphoribosyltransferase deficiency: a previously undescribed genetic defect in man. *J Clin Invest*. 1968;47(10):2281-2289. doi:10.1172/JCI105913

6. Smith AN, Skaug J, Choate KA, et al. Mutations in ATP6N1B, encoding a new kidney vacuolar proton pump 116-kD subunit, cause recessive distal renal tubular acidosis with preserved hearing. *Nat Genet*. 2000;26(1):71-75. doi:10.1038/79208

7. Karet FE, Finberg KE, Nelson RD, et al. Mutations in the gene encoding B1 subunit of H+-ATPase cause renal tubular acidosis with sensorineural deafness. *Nat Genet*. 1999;21(1):84-90. doi:10.1038/5022

8. Gromadzka G, Schmidt HHJ, Genschel J, et al. Frameshift and nonsense mutations in the gene for ATPase7B are associated with severe impairment of copper metabolism and with an early clinical manifestation of Wilson's disease. *Clin Genet*. 2005;68(6):524-532. doi:10.1111/j.1399-0004.2005.00528.x

9. Brennan TM, Landau D, Shalev H, et al. Linkage of infantile Bartter syndrome with sensorineural deafness to chromosome 1p. *Am J Hum Genet*. 1998;62(2):355-361. doi:10.1086/301708

10. Sly WS, Hewett-Emmett D, Whyte MP, Yu YS, Tashian RE. Carbonic anhydrase II deficiency identified as the primary defect in the autosomal recessive syndrome of osteopetrosis with renal tubular acidosis and cerebral calcification. *Proc Natl Acad Sci U S A*. 1983;80(9):2752-2756. doi:10.1073/pnas.80.9.2752

11. Pearce SH, Williamson C, Kifor O, et al. A familial syndrome of hypocalcemia with hypercalciuria due to mutations in the calcium-sensing receptor. *N Engl J Med*. 1996;335(15):1115-1122. doi:10.1056/NEJM199610103351505

12. Lloyd SE, Pearce SHS, Fisher SE, et al. A common molecular basis for three inherited kidney stone diseases. *Nature*. 1996;379(6564):445-449. doi:10.1038/379445a0

13. Schlingmann KP, Konrad M, Jeck N, et al. Salt wasting and deafness resulting from mutations in two chloride channels. *N Engl J Med*. 2004;350(13):1314-1319. doi:10.1056/NEJMoa032843

14. Simon DB, Bindra RS, Mansfield TA, et al. Mutations in the chloride channel gene, CLCNKB, cause Bartter's syndrome type III. *Nat Genet*. 1997;17(2):171-178. doi:10.1038/ng1097-171

15. Simon DB, Lu Y, Choate KA, et al. Paracellin-1, a renal tight junction protein required for paracellular Mg2+ resorption. *Science*. 1999;285(5424):103-106. doi:10.1126/science.285.5424.103

16. Konrad M, Schaller A, Seelow D, et al. Mutations in the tight-junction gene claudin 19 (CLDN19) are associated with renal magnesium wasting, renal failure, and severe ocular involvement. *Am J Hum Genet*. 2006;79(5):949-957. doi:10.1086/508617

17. Town M, Jean G, Cherqui S, et al. A novel gene encoding an integral membrane protein is mutated in nephropathic cystinosis. *Nat Genet*. 1998;18(4):319-324. doi:10.1038/ng0498-319

18. Schlingmann KP, Kaufmann M, Weber S, et al. Mutations in CYP24A1 and idiopathic infantile hypercalcemia. *N Engl J Med*. 2011;365(5):410-421. doi:10.1056/NEJMoa1103864

19. Aponte JL, Sega GA, Hauser LJ, et al. Point mutations in the murine fumarylacetoacetate hydrolase gene: Animal models for the human genetic disorder hereditary tyrosinemia type 1. *Proc Natl Acad Sci U S A*. 2001;98(2):641-645.

20. Jaureguiberry G, De la Dure-Molla M, Parry D, et al. Nephrocalcinosis (enamel renal syndrome) caused by autosomal recessive FAM20A mutations. *Nephron Physiol*. 2012;122(1-2):1-6. doi:10.1159/000349989

21. Yang T, Vidarsson H, Rodrigo-Blomqvist S, Rosengren SS, Enerback S, Smith RJH. Transcriptional control of SLC26A4 is involved in Pendred syndrome and nonsyndromic enlargement of vestibular aqueduct (DFNB4). *Am J Hum Genet*. 2007;80(6):1055-1063. doi:10.1086/518314

22. Seydewitz HH, Matern D. Molecular genetic analysis of 40 patients with glycogen storage disease type Ia: 100% mutation detection rate and 5 novel mutations. *Hum Mutat*. 2000;15(1):115-116. doi:10.1002/(SICI)1098-1004(200001)15:1<115::AID-HUMU23>3.0.CO;2-W

23. Li D, Opas EE, Tuluc F, et al. Autosomal dominant hypoparathyroidism caused by germline mutation in GNA11: phenotypic and molecular characterization. *J Clin Endocrinol Metab*. 2014;99(9):E1774-1783. doi:10.1210/jc.2014-1029

24. Reiss J, Gross-Hardt S, Christensen E, Schmidt P, Mendel RR, Schwarz G. A mutation in the gene for the neurotransmitter receptor-clustering protein gephyrin causes a novel form of molybdenum cofactor deficiency. *Am J Hum Genet*. 2001;68(1):208-213. doi:10.1086/316941

25. Williams HE, Smith LH. L-glyceric aciduria. A new genetic variant of primary hyperoxaluria. *N Engl J Med*. 1968;278(5):233-238. doi:10.1056/NEJM196802012780502

26. Hamilton AJ, Bingham C, McDonald TJ, et al. The HNF4A R76W mutation causes atypical dominant Fanconi syndrome in addition to a β cell phenotype. *J Med Genet*. 2014;51(3):165-169. doi:10.1136/jmedgenet-2013-102066

27. Belostotsky R, Seboun E, Idelson GH, et al. Mutations in DHDPSL are responsible for primary hyperoxaluria type III. *Am J Hum Genet*. 2010;87(3):392-399. doi:10.1016/j.ajhg.2010.07.023

28. Davidson BL, Tarlé SA, Van Antwerp M, et al. Identification of 17 independent mutations responsible for human hypoxanthine-guanine phosphoribosyltransferase (HPRT) deficiency. *Am J Hum Genet*. 1991;48(5):951-958.

29. Simon DB, Karet FE, Rodriguez-Soriano J, et al. Genetic heterogeneity of Bartter's syndrome revealed by mutations in the K+ channel, ROMK. *Nat Genet*. 1996;14(2):152-156. doi:10.1038/ng1096-152

30. Bockenhauer D, Feather S, Stanescu HC, et al. Epilepsy, ataxia, sensorineural deafness, tubulopathy, and KCNJ10 mutations. *N Engl J Med*. 2009;360(19):1960-1970. doi:10.1056/NEJMoa0810276

31. Ichikawa S, Imel EA, Kreiter ML, et al. A homozygous missense mutation in human KLOTHO causes severe tumoral calcinosis. *J Clin Invest*. 2007;117(9):2684-2691. doi:10.1172/JCI31330

32. Kantarci S, Al-Gazali L, Hill RS, et al. Mutations in LRP2, which encodes the multiligand receptor megalin, cause Donnai-Barrow and facio-oculo-acoustico-renal syndromes. *Nat Genet*. 2007;39(8):957-959. doi:10.1038/ng2063

33. Laghmani K, Beck BB, Yang SS, et al. Polyhydramnios, Transient Antenatal Bartter's Syndrome, and MAGED2 Mutations. *N Engl J Med*. 2016;374(19):1853-1863. doi:10.1056/NEJMoa1507629

34. Munroe PB, Olgunturk RO, Fryns JP, et al. Mutations in the gene encoding the human matrix Gla protein cause Keutel syndrome. *Nat Genet*. 1999;21(1):142-144. doi:10.1038/5102

35. Ichida K, Matsumura T, Sakuma R, Hosoya T, Nishino T. Mutation of human molybdenum cofactor sulfurase gene is responsible for classical xanthinuria type II. *Biochem Biophys Res Commun*. 2001;282(5):1194-1200. doi:10.1006/bbrc.2001.4719

36. Shalata A, Mandel H, Reiss J, et al. Localization of a gene for molybdenum cofactor deficiency, on the short arm of chromosome 6, by homozygosity mapping. *Am J Hum Genet*. 1998;63(1):148-154. doi:10.1086/301916

37. Attree O, Olivos IM, Okabe I, et al. The Lowe's oculocerebrorenal syndrome gene encodes a protein highly homologous to inositol polyphosphate-5-phosphatase. *Nature*. 1992;358(6383):239-242. doi:10.1038/358239a0

38. Feske S, Gwack Y, Prakriya M, et al. A mutation in Orai1 causes immune deficiency by abrogating CRAC channel function. *Nature*. 2006;441(7090):179-185. doi:10.1038/nature04702

39. Becker MA, Losman MJ, Wilson J, Simmonds HA. Superactivity of human phosphoribosyl pyrophosphate synthetase due to altered regulation by nucleotide inhibitors and inorganic phosphate. *Biochim Biophys Acta*. 1986;882(2):168-176. doi:10.1016/0304-4165(86)90151-0

40. Enomoto A, Kimura H, Chairoungdua A, et al. Molecular identification of a renal urate anion exchanger that regulates blood urate levels. *Nature*. 2002;417(6887):447-452. doi:10.1038/nature742

41. Gee HY, Jun I, Braun DA, et al. Mutations in SLC26A1 Cause Nephrolithiasis. *Am J Hum Genet*. 2016;98(6):1228-1234. doi:10.1016/j.ajhg.2016.03.026

42. Akagi M, Inui K, Nakajima S, et al. Mutation analysis of two Japanese patients with Fanconi-Bickel syndrome. *J Hum Genet*. 2000;45(1):60-62. doi:10.1007/s100380050013

43. Matsuo H, Chiba T, Nagamori S, et al. Mutations in glucose transporter 9 gene SLC2A9 cause renal hypouricemia. *Am J Hum Genet*. 2008;83(6):744-751. doi:10.1016/j.ajhg.2008.11.001

44. Prié D, Beck L, Friedlander G, Silve C. Sodium-phosphate cotransporters, nephrolithiasis and bone demineralization. *Curr Opin Nephrol Hypertens*. 2004;13(6):675-681. doi:10.1097/00041552-200411000-00015

45. Lorenz-Depiereux B, Benet-Pages A, Eckstein G, et al. Hereditary hypophosphatemic rickets with hypercalciuria is caused by mutations in the sodium-phosphate cotransporter gene SLC34A3. *Am J Hum Genet*. 2006;78(2):193-201. doi:10.1086/499410

46. Calonge MJ, Gasparini P, Chillarón J, et al. Cystinuria caused by mutations in rBAT, a gene involved in the transport of cystine. *Nat Genet*. 1994;6(4):420-425. doi:10.1038/ng0494-420

47. Bruce LJ, Cope DL, Jones GK, et al. Familial distal renal tubular acidosis is associated with mutations in the red cell anion exchanger (Band 3, AE1) gene. *J Clin Invest*. 1997;100(7):1693-1707. doi:10.1172/JCI119694

48. Turk E, Zabel B, Mundlos S, Dyer J, Wright EM. Glucose/galactose malabsorption caused by a defect in the Na+/glucose cotransporter. *Nature*. 1991;350(6316):354-356. doi:10.1038/350354a0

49. Feliubadaló L, Font M, Purroy J, et al. Non-type I cystinuria caused by mutations in SLC7A9, encoding a subunit (bo,+AT) of rBAT. *Nat Genet*. 1999;23(1):52-57. doi:10.1038/12652

50. Karim Z, Gérard B, Bakouh N, et al. NHERF1 mutations and responsiveness of renal parathyroid hormone. *N Engl J Med*. 2008;359(11):1128-1135. doi:10.1056/NEJMoa0802836

51. Scott P, Ouimet D, Valiquette L, et al. Suggestive evidence for a susceptibility gene near the vitamin D receptor locus in idiopathic calcium stone formation. *J Am Soc Nephrol*. 1999;10(5):1007-1013. doi:10.1681/ASN.V1051007

52. El-Sayed W, Parry DA, Shore RC, et al. Mutations in the beta propeller WDR72 cause autosomal-recessive hypomaturation amelogenesis imperfecta. *Am J Hum Genet*. 2009;85(5):699-705. doi:10.1016/j.ajhg.2009.09.014

53. Dent CE, Philpot GR. Xanthinuria, an inborn error (or deviation) of metabolism. *Lancet*. 1954;266(6804):182-185. doi:10.1016/s0140-6736(54)91257-x
