## supplementary table 2 for "Beyond the Kidney: Extra-Renal Manifestations of Monogenic Nephrolithiasis and Their Significance"

| | Phenotype Category (n=24) | | | | | | | | | | | | | | | | | | | | | | | | |
|---|---|---|---|---|---|---|---|---|---|---|---|---|---|---|---|---|---|---|---|---|---|---|---|---|---|
| ***Gene-Associated Phenotype (n=55)*** | **Growth** | **Respiratory** | **Cardiovascular** | **Endocrine** | **Neurologic** | **Nose** | **Mouth** | **Teeth** | **Eyes** | **Neck** | **Face** | **Ears** | **Head** | **Chest** | **Abdomen/ Gastrointestinal** | **Muscle** | **Skin** | **Skeletal** | **Hematology** | **Immunology** | **Metabolic Features** | **Prenatal Manifestations** | **Voice** | **Neoplasia** | **Number of phenotype categories involved per GAP (%)** |
| ***ADCY10*** | N | N | N | N | N | N | N | N | N | N | N | N | N | N | N | N | N | N | N | N | N | N | N | N | 1 (4%) |
| ***AGXT*** | N | N | Y | N | Y | N | N | Y | Y | N | N | N | N | N | N | N | Y | Y | N | N | Y | N | N | N | 8 (33%) |
| ***ALDOB*** | Y | N | N | N | Y | N | N | Y | N | N | N | N | N | N | Y | N | Y | N | N | N | Y | N | N | N | 7 (29%) |
| ***ALPL*** | Y | Y | N | N | Y | N | N | Y | Y | N | Y | N | Y | Y | Y | N | Y | Y | Y | N | N | Y | Y | N | 14 (58%) |
| ***APRT*** | N | N | N | N | N | N | N | N | N | N | N | N | N | N | N | N | N | N | N | N | N | N | N | N | 1 (4%) |
| ***ATP6V0A4*** | N | N | N | N | N | N | N | N | N | N | N | Y | N | N | N | N | N | Y | N | N | Y | N | N | N | 4 (17%) |
| ***ATP6V1B1*** | Y | N | N | N | N | N | N | N | N | N | N | Y | N | N | Y | N | N | Y | N | N | Y | N | N | N | 6 (25%) |
| ***ATP7B*** | N | N | N | Y | Y | N | N | N | Y | N | N | N | N | N | Y | N | N | Y | Y | N | N | N | N | N | 7 (29%) |
| ***BSND*** | N | N | N | N | N | N | N | N | N | N | N | N | N | N | N | N | N | N | N | N | N | N | N | N | 1 (4%) |
| ***CA2*** | Y | N | N | N | Y | N | N | Y | N | N | N | Y | N | N | N | Y | N | Y | Y | N | Y | N | N | N | 8 (33%) |
| ***CASR*** | Y | Y | N | Y | Y | N | N | N | N | N | N | N | N | Y | Y | Y | N | Y | Y | N | N | N | N | N | 10 (42%) |
| ***CLCN5*** | Y | N | N | N | N | N | N | N | N | N | N | N | N | N | N | N | N | Y | N | N | N | N | N | N | 3 (13%) |
| ***CLCNKA*** | Y | N | N | Y | Y | N | N | N | N | N | N | Y | N | N | N | Y | N | N | N | N | N | Y | N | N | 7 (29%) |
| ***CLCNKB*** | Y | N | Y | Y | Y | N | N | N | Y | N | N | Y | N | N | N | Y | N | N | N | N | Y | Y | N | N | 10 (42%) |
| ***CLDN16*** | Y | N | N | N | Y | N | N | N | Y | N | N | N | N | N | Y | Y | N | N | N | N | Y | N | N | N | 7 (29%) |
| ***CLDN19*** | N | N | N | N | N | N | N | Y | Y | N | N | N | N | N | N | N | N | N | N | N | N | N | N | N | 3 (13%) |
| ***CTNS*** | Y | N | N | Y | Y | N | N | N | Y | N | Y | N | N | Y | Y | Y | Y | Y | N | N | Y | N | Y | N | 13 (54%) |
| ***CYP24A1*** | Y | N | N | N | Y | N | N | N | N | N | N | N | N | N | Y | Y | N | N | N | N | Y | N | N | N | 6 (25%) |
| ***FAH*** | Y | N | Y | N | Y | N | N | N | N | N | N | N | N | N | Y | Y | N | Y | Y | N | Y | N | N | Y | 10 (42%) |
| ***FAM20A*** | N | N | N | N | N | N | N | Y | N | N | N | N | N | N | N | N | N | N | N | N | Y | N | N | N | 3 (13%) |
| ***FOXI1*** | N | N | N | N | N | N | N | N | N | N | N | Y | N | N | N | N | N | N | N | N | N | N | N | N | 1 (4%) |
| ***G6PC*** | Y | N | Y | N | N | N | N | N | Y | N | Y | N | N | N | Y | Y | Y | Y | Y | N | Y | N | N | N | 11 (46%) |
| ***GNA11*** | Y | N | N | Y | Y | N | N | N | N | N | N | N | N | N | N | Y | N | N | N | N | N | N | N | N | 4 (17%) |
| ***GPHN*** | N | N | N | N | Y | N | N | N | Y | N | N | N | N | N | Y | Y | N | N | N | N | N | N | N | N | 4 (17%) |
| ***GRHPR*** | N | N | N | N | N | N | N | N | N | N | N | N | N | N | N | N | N | N | N | N | N | N | N | N | 1 (4%) |
| ***HNF4A*** | Y | N | N | Y | N | N | N | N | N | N | N | N | N | N | Y | N | N | Y | N | N | N | N | N | N | 5 (21%) |
| ***HOGA1*** | N | N | N | N | N | N | N | N | N | N | N | N | N | N | N | N | N | N | N | N | N | N | N | N | 1 (4%) |
| ***HPRT1*** | Y | N | N | N | Y | N | N | N | N | N | N | N | N | N | Y | N | Y | Y | Y | N | N | N | N | N | 7 (29%) |
| ***KCNJ1*** | Y | N | Y | Y | Y | N | N | N | Y | N | Y | Y | Y | N | Y | Y | N | Y | Y | N | N | Y | N | N | 14 (58%) |
| ***KCNJ10*** | Y | N | Y | N | Y | N | N | N | N | N | N | Y | N | N | Y | N | N | N | N | N | Y | N | N | N | 7 (29%) |
| ***KL*** | N | N | Y | Y | Y | N | N | Y | N | N | N | N | N | N | N | N | N | Y | N | N | N | N | N | N | 5 (21%) |
| ***LRP2*** | Y | Y | Y | N | Y | Y | N | N | Y | N | Y | Y | Y | Y | Y | N | Y | Y | N | N | N | N | N | N | 14 (58%) |
| ***MAGED2*** | N | N | N | N | N | N | N | N | N | N | N | N | N | N | N | N | N | N | N | N | N | Y | N | N | 2 (8%) |
| ***MGP*** | Y | Y | Y | N | Y | Y | N | N | N | N | Y | Y | N | Y | N | N | Y | Y | N | N | N | Y | Y | N | 12 (50%) |
| ***MOCOS*** | N | N | N | N | N | N | N | N | N | N | N | N | N | N | N | Y | N | N | N | N | N | N | N | N | 1 (4%) |
| ***MOCS1*** | Y | N | N | N | Y | Y | Y | N | Y | N | Y | N | Y | N | Y | Y | N | Y | N | N | N | N | N | N | 10 (42%) |
| ***MOCS2*** | Y | N | N | N | Y | Y | Y | N | Y | N | Y | N | Y | N | Y | Y | N | Y | N | N | N | N | N | N | 10 (42%) |
| ***OCRL*** | Y | N | N | N | Y | N | N | Y | Y | N | N | N | N | N | Y | N | Y | Y | N | N | Y | Y | N | N | 10 (42%) |
| ***ORAI1*** | Y | Y | N | N | Y | N | Y | Y | Y | Y | N | N | N | N | N | Y | Y | Y | N | Y | Y | N | N | N | 12 (50%) |
| ***PRPS1*** | N | N | N | N | Y | N | N | N | N | N | N | Y | N | N | N | N | N | Y | N | N | Y | N | N | N | 5 (21%) |
| ***SLC12A1*** | Y | N | Y | Y | Y | N | N | N | N | N | N | N | N | N | Y | Y | N | Y | N | N | Y | Y | N | N | 10 (42%) |
| ***SLC22A12*** | N | N | N | N | N | N | N | N | N | N | N | N | N | N | N | N | N | N | N | N | N | N | N | N | 1 (4%) |
| ***SLC26A1*** | N | N | N | N | N | N | N | N | N | N | N | N | N | N | N | N | N | N | N | N | N | N | N | N | 1 (4%) |
| ***SLC2A2*** | Y | N | N | N | Y | N | N | N | N | N | N | N | N | N | Y | Y | N | Y | N | N | Y | N | N | N | 7 (29%) |
| ***SLC2A9*** | N | N | N | N | N | N | N | N | N | N | N | N | N | N | N | N | N | N | N | N | N | N | N | N | 1 (4%) |
| ***SLC34A1*** | Y | N | N | Y | N | N | N | N | N | N | N | N | N | N | N | Y | N | Y | N | N | N | N | N | N | 5 (21%) |
| ***SLC34A3*** | Y | N | N | N | N | N | N | N | N | N | N | N | Y | Y | Y | Y | N | Y | N | N | N | N | N | N | 7 (29%) |
| ***SLC3A1*** | N | N | N | N | N | N | N | N | N | N | N | N | N | N | N | N | N | N | N | N | N | N | N | N | 1 (4%) |
| ***SLC4A1*** | Y | N | N | N | Y | N | N | N | N | N | N | N | N | N | Y | N | Y | Y | Y | N | Y | N | N | N | 8 (33%) |
| ***SLC5A1*** | Y | N | N | N | N | N | N | N | N | N | N | N | N | N | Y | N | N | N | N | N | Y | N | N | N | 3 (13%) |
| ***SLC7A9*** | N | N | N | N | N | N | N | N | N | N | N | N | N | N | N | N | N | N | N | N | N | N | N | N | 1 (4%) |
| ***SLC9A3R1*** | N | N | N | N | N | N | N | N | N | N | N | N | N | N | N | N | N | Y | N | N | N | N | N | N | 2 (8%) |
| ***VDR*** | Y | N | N | Y | Y | N | N | Y | N | N | N | Y | Y | Y | Y | Y | Y | Y | N | N | N | N | N | N | 11 (46%) |
| ***WDR72*** | N | N | N | N | N | N | N | Y | N | N | N | N | N | N | N | N | N | N | N | N | N | N | N | N | 1 (4%) |
| ***XDH*** | N | N | N | N | N | N | N | N | N | N | N | N | N | N | N | Y | N | N | N | N | N | N | N | N | 2 (8%) |
| **Number of GAPs (%)** | 31 (56%) | 5 (9%) | 10 (18%) | 12 (22%) | 29 (53%) | 4 (7%) | 3 (5%) | 11 (20%) | 15 (27%) | 1 (2%) | 8 (15%) | 12 (22%) | 7 (13%) | 7 (13%) | 25 (45%) | 22 (40%) | 12 (22%) | 29 (53%) | 9 (16%) | 1 (2%) | 20 (36%) | 8 (15%) | 3 (5%) | 1 (2%) | |
| **P-value** | <0.001 | 0.16 | 0.99 | 0.63 | <0.001 | 0.08 | 0.04 | 0.80 | 0.26 | <0.001 | 0.60 | 0.60 | 0.43 | 0.43 | <0.001 | 0.01 | 0.60 | <0.001 | 0.80 | <0.001 | 0.03 | 0.60 | 0.03 | <0.001 | |

**Supplementary Table 2.**

**A detailed breakdown of each gene's impact on every phenotype category, including the number of categories affected by each gene and the number of genes that impact each category. Additionally, the results of the chi-square test for each category are included.**

**Y: Yes; involved**

**N: No; not involved**

**GAP: gene-associated phenotype**
